# Effect of Maximum Repetition of Pelvic Floor Stabilization Exercise in Stress Urinary Incontinence

**DOI:** 10.1101/2021.11.02.21265704

**Authors:** Iren Khatun, Mohammad Anwar Hossain, K M Amran Hossain, Nadia Afrin Urme

**Affiliations:** Department of physiotherapy, Bangladesh Health Professions Institute (BHPI), Dhaka, Bangladesh; Department of Physiotherapy, Centre for the Rehabilitation of the Paralysed (CRP), Dhaka, Bangladesh

**Keywords:** Stress Urinary Incontinence, Physiotherapy, exercise, Maximum repetition

## Abstract

**Background:** Stress urinary incontinence (SUI) in females is a common gynecological issue that impedes lifestyle. Exercise had a significant effect; however, studies did not determine the exercise frequency and intensity for pelvic floor stabilization in stress urinary incontinence.

**Aim:** The aim of the study is to determine if maximum repetition of pelvic stabilization exercise impacts the management of stress urinary incontinence in females.

**Methodology:** One arm quasi-experimental study design was used. 40 patients having SUI and associated musculoskeletal complaints were recruited from the outpatient unit of Physiotherapy department of the Centre for the Rehabilitation of the Paralysed (CRP), Bangladesh. The study was conducted over 4weeks. Outcome measurement was included pelvic floor and abdominal muscle strength, endurance, and incontinence measurement.

**Result:** Pelvic floor muscle and abdominal strength, and endurance had a positive and significant result in maximum repetition (P .001). Pelvic floor strength has been significantly improvement in week 2 (P .001), and week 3 (P .01). Interference in activities (P .003), and ICIQ total (P .001) had improvement but majority of the improvement was noted in weeks 2-3. There was a significant improvement in the frequency of urine leakage in the first week (P .001), and week 3 (P .005) and week 4 (P .001).

**Conclusion:** Pelvic floor exercise with increasing repetition is an effective approach to improve stress urinary incontinence in females. The study had a significant impact on incontinence frequency, amount, and associated quality of life for women with stress urinary incontinence with pelvic floor exercise with maximum repetition.

## 1. Background

Stress urinary incontinence (SUI), characterized as “objection of compulsory loss of urine on exertion or actual effort (e.g., brandishing exercises) or on wheezing or hacking.” Missing are the manifestations of an overactive bladder criticalness, nocturia, and enuresis [1]. Urinary incontinence (UI) is a common problem among adults living in the community. Its incidence increases with age and it is more frequent in women, being particularly common amongst elderly women in residential care. Besides childbirth, smoking, chronic bronchitis, and obesity also acts as a risks factor for this. Estimates of the prevalence of urinary incontinence in women vary from 10% up to 40%. Irrespective old enough, 15% to 30% of women are influenced by urinary incontinence in all aspects of their lives physical, mental and social with ensuing weakening in personal satisfaction [2,3].

Pelvic floor muscle training (PFMT) consists of a programme of repeated contractions and relaxations of the pelvic floor muscles taught and supervised by a health professional. PFMT is the most commonly used physical therapy for women with stress urinary incontinence (SUI) [4]. It offers a possible reprieve from urinary incontinence. This conservative therapy appears to have no significant side effects and enables improvement in symptoms; it can therefore be considered as a first choice of treatment for urinary incontinence in women [5]. Many studies found significant impact of pelvic floor stabilization on stress incontinence, but they did not determine the impact of maximum repetition of pelvic floor stabilization exercise. Therefore, Primary objective of this research is to determine the maximum repetition of pelvic stabilization exercise upon the management of stress urinary incontinence in females. Secondary objective is to (1) explore socio-demographics related to SUI, (2) observe the impact of maximum repetition of pelvic floor stabilization exercise upon pelvic floor strength & endurance, transverse abdominis strength and functional disability in stress urinary incontinence patients, (3) observe the changes as per repeated measurement weekly in 4 weeks. This study hypothesizes that there is a positive effect of Maximum repetition of Pelvic floor stabilization Exercise in Stress Urinary Incontinence compared to care.

## 2. Method

### 2.1 Study design

A one arm pretest and repeated posttest design of Quasi-experimental study was conducted to find out the effectiveness of maximum repetitions of pelvic floor stabilization exercise for treating stress urinary incontinence.

### 2.2 Study Participants and Settings

Participants were recruited from the musculoskeletal department, Centre for the Rehabilitation of the Paralysed (CRP), Bangladesh. Patient came to an outpatient unit with low back pain and stress urinary incontinence (from 1^st^November 2019 to 20^th^February 2020) has been chosen as the study population. Primarily 58 subjects have been screened with Low Back Pain and urinary incontinence and from which 40 patients have been confirmed by consultant Physiotherapist based on the eligibility criteria. Eligibility criteria were- (1) diagnosed case of Stress urinary incontinence according to ICD 10 [6]; (2) Age 30-75 years of age [7]; (3) Both prime or multipara [7]; (4) Any surgery in the genito-urinary tract [8]; (5) Patient with other musculoskeletal complaints (LBP/ arthritis) as they attended the Musculoskeletal Unit of the Physiotherapy Department. Participants were excluded- (1) having surgery for incontinence; (2) Mixed incontinence as per ICD 10 [9]; (3) Carcinoma or critically ill patients and UTI or genitor-urinary infections.

### 2.3 Sampling Technique

Total 58 participants were assessed for eligibility. Among them 40 participants meet the criteria and gave consent to participate in the study (Figure-1). The study samples have been drawn from the population through a hospital-based randomization process. As these patients accomplished this CRP haphazardly without the decision of a CRP expert or the specialist’s decision, so they might be considered as a random example.

**Figure 1:**
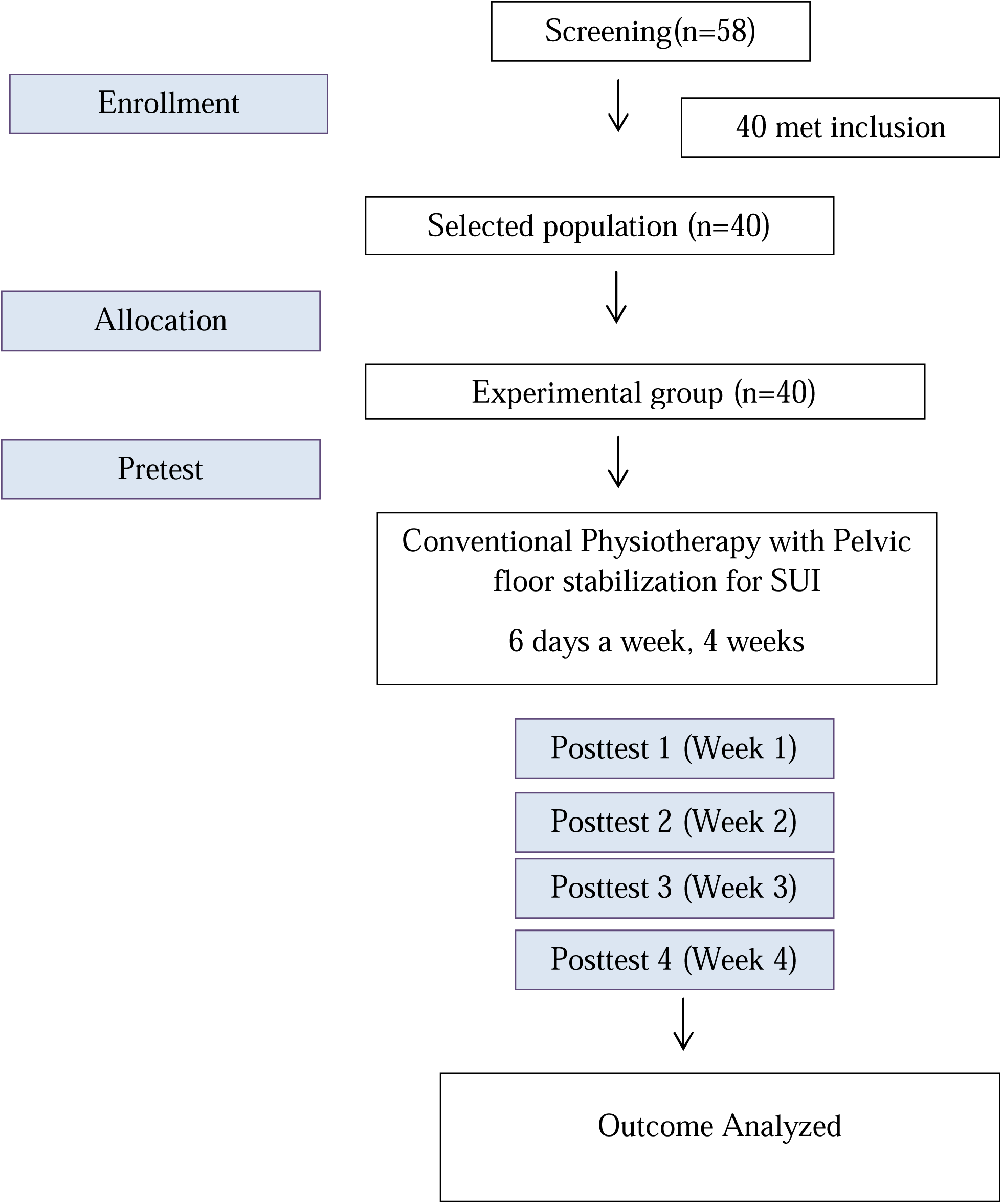
The Template for Intervention Description and Replication (TIDieR)flow chart.

**Figure 2:**
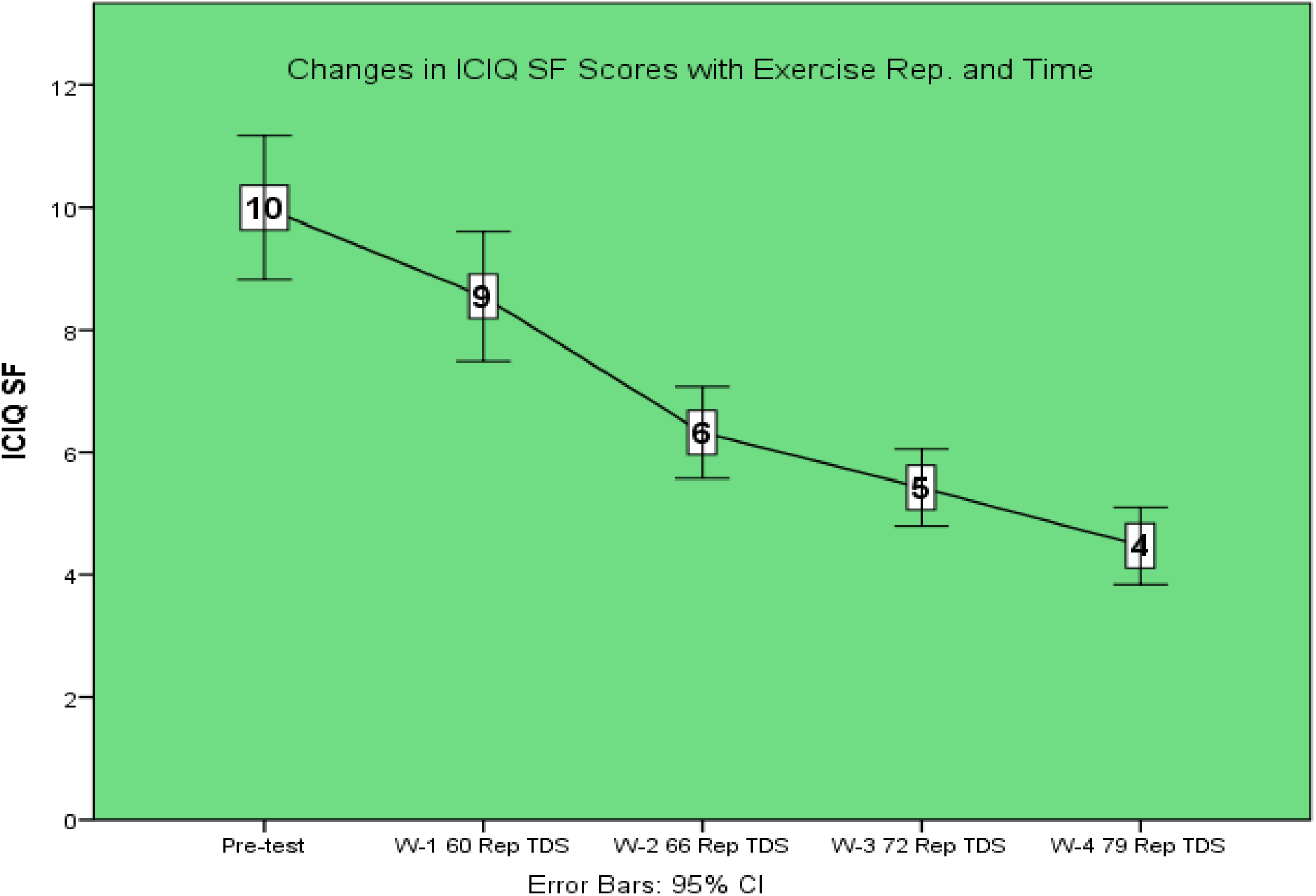
Changes in ICIQSF

### 2.4. Intervention

The patients were received McKenzie treatment for Low Back pain. The McKenzie therapy was included, repeated movements typically including flexion in lying or standing; extension in lying or standing; and lateral movements of either side gliding or rotation and manipulative approach to lumbar spine segments [10]. Patients were performed those movements at therapy sessions and at home [11]. The repeated movements of McKenzie therapy were prescribed as 10 repetitions of directed movements, 2-3 hourly in 14 hours of a day and for 4 weeks. McKenzie therapies were performed by physiotherapists for weeks. Besides they were receive manual passive stretching exercise for lumbo-pelvic muscles for 5-7 repetitions per muscle with 10-15 seconds hold performed twice a day for 2 weeks and graded oscillatory mobilization in Maitland concept in 5-7 minutes, 35-40 oscillation per minutes or static segmental mobilizations in Maitland concept for 35-50 second hold for 5-7 times in lumbar spine for 6 sessions in 2 weeks In addition, they were received analgesics and hot compression in lower back for 10 minutes for 2 weeks, stabilization exercises of lumbo-pelvic segment accompanied with a booklet indicating the proper way to do different activities and lifestyles habits for 4 weeks [12] Besides treatment for low back pain, patients received the pelvic floor exercise three times a day (dx.doi.org/10.17504/protocols.io.by4npyve)

#### Intervention provider

Graduate physiotherapists who are expert in treatment of musculoskeletal patients have been involved in treatment of patients. Treatment providers had the experience of more than 2years, in the aspect of musculoskeletal physiotherapy. Researchers arranged service training to share the information. Practical demonstration involved training on the intervention including procedure, dose, intensity, frequency, repetition and patient position. In addition, the types, dose repetition, duration of conventional care including manual therapy, exercise therapy and electrotherapy has been taken from head of Physiotherapy department, Centre for the Rehabilitation of the Paralyzed (CRP).

### 2.5. Intervention outcome

#### Incontinence Measurement

Urinary incontinence was measured by using International Consultation on Incontinence Questionnaire Short Form (ICIQ SF). It is a patient reported questionnaire that is valid and reliable for incontinence measurement. There are 4 questions for symptoms in the last 1 month, one has the sum 3,4,5 can be called as actual score, where first 2 items are demographic questions. Final score is un-scored and self-diagnostic features. Scoring enabled by 0-21, where o is the least result and 21 is the highest. The question is valid for 18-64 years and 65 years or above aged respondents. The test-retest reliability is .74, correlation coefficients were .93 and .96 respectively [13,14,15].

#### Manual Muscle Testing

Pelvic floor muscle strength was measured by manual muscle testing. Laycock established the Modified Oxford Grading System to measure the strength of pelvic floor muscle. It is a six-point scale and measured by vaginal palpation. Points are included, 0= no contraction, 1=flicker, 2=weak, 3=moderate, 4= good (with lift), 5=strong. It is a commonly used and accepted tools by physiotherapists due to cost effectiveness. The tools show high inter-rater reliability (κ =.33, 95% CI interval .09-.57) [16].

### 2.6. Data collection process

Data collection process was conducted through screening the patient for eligibility of participation, intervention and outcome measurement. Screening has been carried out by a physiotherapist. Then 40 subjects have been screened with SUI which has been confirmed by consultant Physiotherapist based on the inclusion criteria. Consent was taken from the participation before starting treatment. A baseline assessment has been done then according to the previous discussed protocol or flowchart that has been provided. After baseline determination, a post-test is performed every week. The posttest of the last patient was completed in mid-march 2020 before the lock down due to COVID-19 pandemic started. No blinding or masking done for the study.

### 2.7. Data analysis

Statistical analysis has been performed by using the statistical package for social science (SPSS) version 20. Microsoft Excel was used to design pie charts, bar charts, and linear line diagrams. Descriptive statistics has been performed as per the nature of data. For parametric data mean and standard deviation has been calculated and for non-parametric data frequency distribution has been checked. Inferential statistics performed as Repeated Measure ANOVA for parametric data to analyze the changes as per weeks, also a week wise comparison enabled using paired t-test. Friedman’s ANOVA has been used instead of Repeated Measure ANOVA for non-parametric data and a post-HOC analysis by Wilcoxon test week to week. The alpha value has been set P<.05 and in Post HOC test value calculated as P<.0125

### 2.8. Quality control and confirmation

The specialist had enough learning in the assigned examination, henceforth the investigation zone also, underneath issues had been acutely investigated by him. The arrangement of the study was simply basic; accordingly, it empowered a complete answer. The trial was created by the review of literature; pursue the universal acknowledged trial and companion explored for dependable poll. The examiner endeavored to keep away from choice predisposition because of carefully kept up incorporation and exclusion criteria.

The examination stayed away from strife with the determination of the members. The information was gathered by an experienced physiotherapist who distinguished lumbar plate prolapsed patients as members. The data has been collected by a separate data collector employed for the study.

### 2.9 Ethical issues

The whole process of this research project has been done by following the national guidelines of Bangladesh Medical Research Council (BMRC) and World Health Organization (WHO) Research guidelines. A written approval from the Institutional Review Board (IRB) of Bangladesh Health Professions Institute has been obtained. After getting permission from the IRB of BHPI, trial registration **(CTRI/2020/08/027107)** was taken from the Indian trial registry board. Then the data collection started. For data collection, a separate approval from the Head-Department of Physiotherapy, CRP has been taken. During the data collection procedure-written consent has been taken from the patients. Every participant had the right to proceed or withdraw from the study anytime.

## 3. RESULTS

### 3.1 Baseline characteristics of the participants

#### Demographic and incontinence related information

40 participants completed the study. The mean age of participants was 48.32±12.04 years, with the minimum age 30 years and maximum 75 years. 32.5% (n=13) participants were from 30-40 years, 42.5% (n=17) were from 41-55years, 20% (n=8) from 56-70years, and 5% (n=2) respondents aged more than 70 years. The mean height was 156.5±5.2 cm. The mean weight was 67±8.4 Kg. The BMI mean was 27.4±3, the majority of the respondents were overweight. 10% of the respondents were prime Para, 40% had 2 children, 17.5% had 3 children, 20% had 4 children, 5% had 5 children, 2.5% had 6 children and 5% respondents had 8 children. Rural residents were 32% and from urban areas 68%. There were several occupational women including -housewife (82.5%), teacher (5%) and other service holder (12.5%).17.5% were Illiterate, majority were primary educated (70%), and 12.5% were graduates. Besides, from the participants 35% were suffering from Diabetes Mellitus, 47.5% of people had hypertension, and 17.5% had Diabetes, hypertension and multiple comorbidities. 45% had a gynecological surgery not related with bladder.

#### Strength and endurance of pelvic floor and abdominal muscle

25% of the population (n=10) had Pelvic floor strength 0 in Manual Muscle test, 57.5% had strength 1 and 17.5% had 2 out of 5. Similarly, 4% had Abdominal muscle strength 1, 52.5% had 2, 35% had strength 3 and 2.5% had abdominal muscle strength 4 out of 5. Pelvic floor endurance varied from minimum 0 second to 56 seconds. The mean was 14.45±10.4 seconds. Abdominal muscle endurance varied from a minimum 7 seconds to maximum 47 seconds, the mean was 19.28±10.5 seconds during baseline assessment.

#### Frequency of leak urine

25% (n=10) stated they had a leak in urine once a week, 33.5% said they leak urine 2-3 times a week, 15% said they had leaking urine every day, 35% stated they leak urine several times a day and 2.5% said they leak urine frequently.

#### Severity of Urine leak and Interference

The amount of leaking urine varies from Small Amount 45% and large amount (55%). The respondents stated their incontinence interference with their daily living in (0-10 scale) as mean 3.85±2.02. The ICIQ SF total from 5-18 scale was 10±3.6.

#### Activities provoke urine leakage

Majority of the patients stated they leak urine during cough or sneeze (80%), during physical activity (12.5%) and after urination (7.5%).

### 3.2. Changes with number of repetitions

Repeated measure ANOVA was used to see the impact of increasing repetition number. Pelvic floor muscle and abdominal muscle strength has a positive and statistically significant (F= .557, p= .001; F=.130, P= .001) result with increasing number of repetitions. Similarly, Pelvic floor muscle and abdominal muscle endurance has a positive and statistically significant (F=.158’ P=.002; F=.173, P=.001) result in repetition. Overall Interference in activities due to leaking and ICIQ score was also calculated (F=.323, P=.003; F= .214 and P=.001) **(Table-1)**.

**Table-1:**
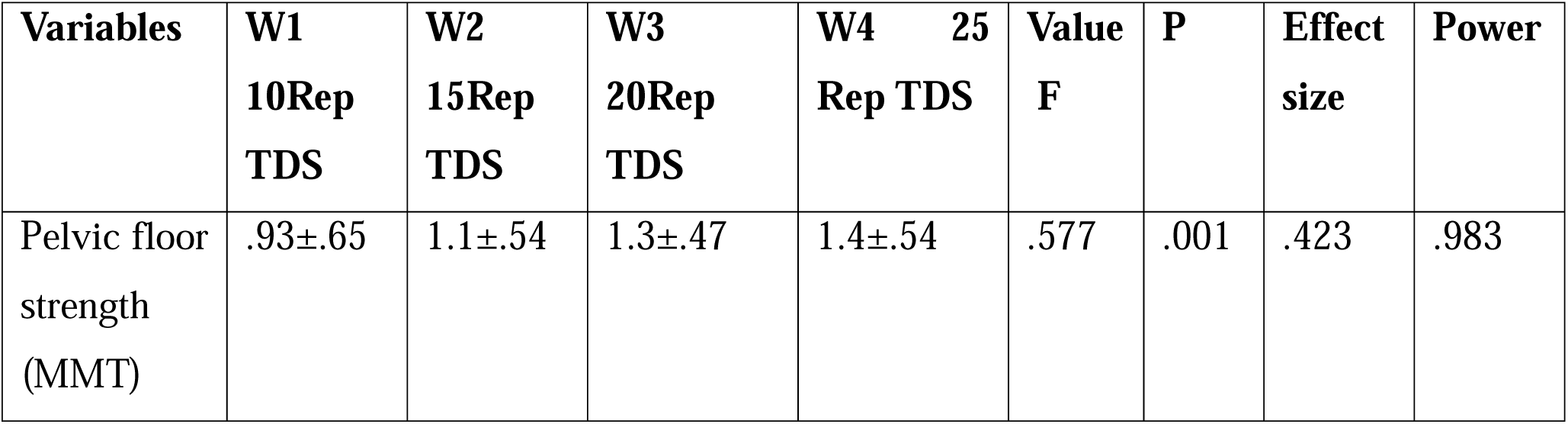
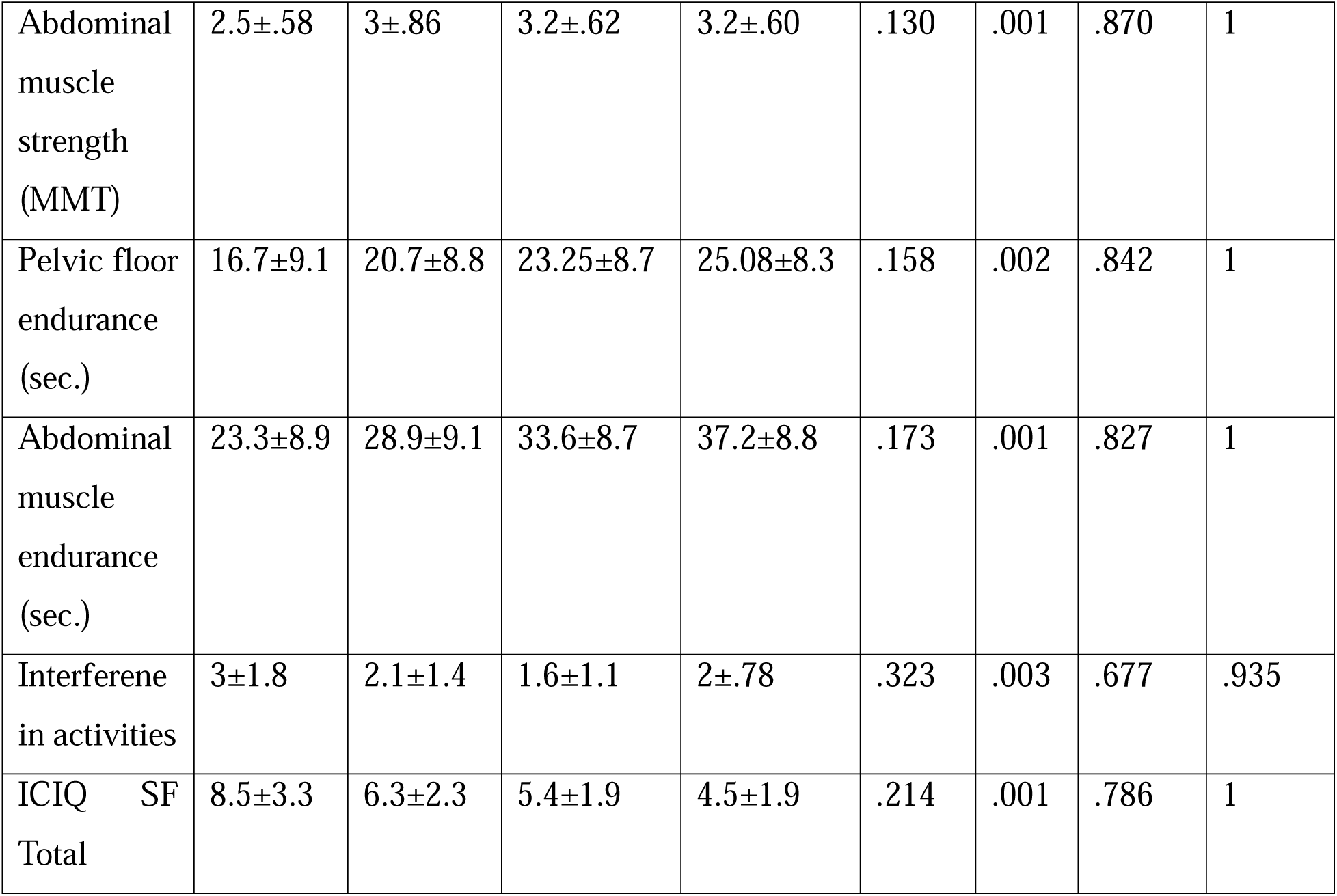
Changes with number of repetitions

| **Variables** | **W1**<br>**10Rep**<br>**TDS** | **W2**<br>**15Rep**<br>**TDS** | **W3**<br>**20Rep**<br>**TDS** | **W4 25**<br>**Rep TDS** | **Value**<br>**F** | **P** | **Effect**<br>**size** | **Power** |
| --- | --- | --- | --- | --- | --- | --- | --- | --- |
| Pelvic floor<br>strength<br>(MMT) | .93±.65 | 1.1±.54 | 1.3±.47 | 1.4±.54 | .577 | .001 | .423 | .983 |
| Abdominal muscle strength (MMT) | 2.5±.58 | 3±.86 | 3.2±.62 | 3.2±.60 | .130 | .001 | .870 | 1 |
| Pelvic floor endurance (sec.) | 16.7±9.1 | 20.7±8.8 | 23.25±8.7 | 25.08±8.3 | .158 | .002 | .842 | 1 |
| Abdominal muscle endurance (sec.) | 23.3±8.9 | 28.9±9.1 | 33.6±8.7 | 37.2±8.8 | .173 | .001 | .827 | 1 |
| Interference in activities | 3±1.8 | 2.1±1.4 | 1.6±1.1 | 2±.78 | .323 | .003 | .677 | .935 |
| ICIQ SF Total | 8.5±3.3 | 6.3±2.3 | 5.4±1.9 | 4.5±1.9 | .214 | .001 | .786 | 1 |

### 3.3. Changes as per week

Week wise changes was measured by using paired t-test Pelvic floor strength has shown statistically significant improvement in week 2 (P= .001), and week 3 (P=.01). Similar improvement noted in week 2 (P=.001) and in week 3 (P=.002). Pelvic floor endurance has been statistically significant improvement in week 2 (P .001), and week 3 (P .01). Similar improvement noted in week 2 (P .001) and in week 3 (P .002). Week wise comparison reveals statistically significant improvement of interference in week 2 (P .001), and week 3 (P .003). Similar improvement noted in ICIQ total from week 1 (P .001), week 3 (P .001) and in week 3 (P .001) **(Table-2)**.

**Table-2:** Changes as per week

**Table-1:** Changes with number of repetitions
|  | Pretest to W-1 |  |  | W-1 to W-2 |  |  | W-2 to W-3 |  |  | W-3 to W-4 |  |  |
| --- | --- | --- | --- | --- | --- | --- | --- | --- | --- | --- | --- | --- |
|  | Mean diff. | t | P | Mean diff. | T | P | Mean diff. | t | P | Mean diff. | t | P |
| Pelvic floor strength | .03±.2 | .03 | 1 | -.25±.4 | -<br>3.6 | .001 | -.15±.4 | -<br>2.6 | .01 | -.07±.3 | -<br>1.7 | .08 |
| Abdominal muscle strength | -.07±.4 | -<br>1.4 | .18 | -.6±.5 | -<br>7.6 | .001 | -.23±.4 | -<br>3.4 | .002 | -.05±.3 | -1 | .323 |
| Pelvic floor endurance (sec.) | -<br>2.3±3.6 | -<br>4.1 | .001 | -<br>3.9±3.6 | -<br>6.7 | .001 | -<br>2.5±2.5 | -<br>6.4 | .001 | -<br>1.8±3.3 | -<br>3.4 | .002 |
| Abdominal muscle endurance (sec.) | -4±3.5 | -<br>7.2 | .001 | -<br>5.6±4.2 | -<br>8.3 | .001 | -<br>4.6±4.9 | -6 | .001 | -<br>3.5±3.1 | -<br>7.1 | .001 |
| Interference in activities | .8±1 | 4.2 | .06 | .95±1.3 | 4.6 | .001 | .47±.9 | 3.1 | .003 | -<br>.37±1.2 | -<br>1.8 | .07 |
| ICIQ SF Total | 1.4±1.8 | 4.9 | .001 | 2.2±2 | 6.7 | .001 | .9±1.6 | 3.4 | .001 | .95±1.2 | 5.2 | .001 |

### 3.4. Improve in Urine Leakage

The frequency of urine leakage and provocation of activities has been analyzed by non-parametric Friedman’s ANOVA that is alternative to repeated measure ANOVA. There were statistically significant results in “how often leaks urine” with X^2^ 84.9 and significant value .001; and amount of urine leaks X^2^ 95 with significant value .003 **(Table-3)**.

**Table -3:** Improve of urine leakage

**Table-2:** Changes as per week
| Variables | Pre-test | W1<br>10Rep<br>TDS | W2<br>15Rep<br>TDS | W3<br>20Rep<br>TDS | W4 25<br>Rep TDS | X<sup>2</sup> -<br>Value | df | P |
| --- | --- | --- | --- | --- | --- | --- | --- | --- |
| How Often leaks | 3.98 | 3.84 | 2.79 | 2.42 | 1.99 | 84.9 | 4 | .001 |
| Urine |  |  |  |  |  |  |  |  |
| When Urine<br>leaks | 3.05 | 3.05 | 3.05 | 2.99 | 2.86 | 4.0 | 4 | .40 |
| Amount of urine<br>leaks | 3.91 | 3.53 | 3.14 | 2.91 | 1.51 | 95 | 4 | .003 |

In week wise comparison, Wilcoxon test has been employed instead of paired t test. Hence there was significant improvement in frequency of urine leakage in the first week (P .001), and week 3 (.005) and week 4 (.001). The time of urine leakage and amount of leakage in every week **(Table-4)**.

**Table-4:**
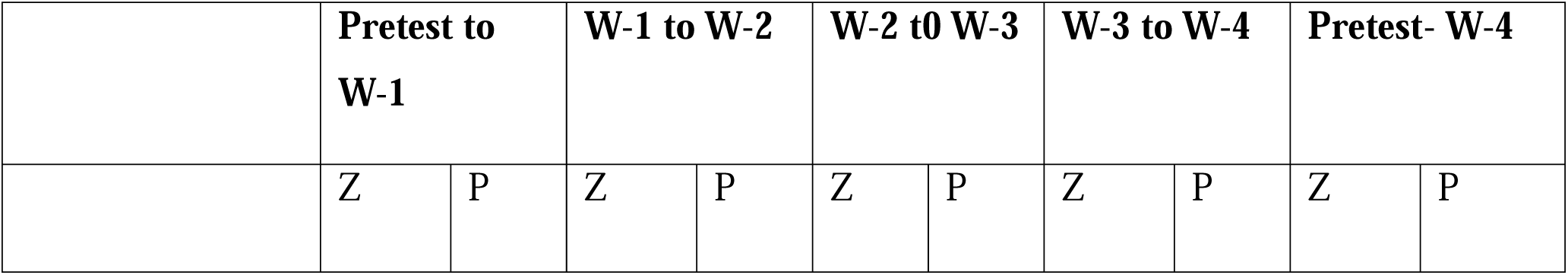
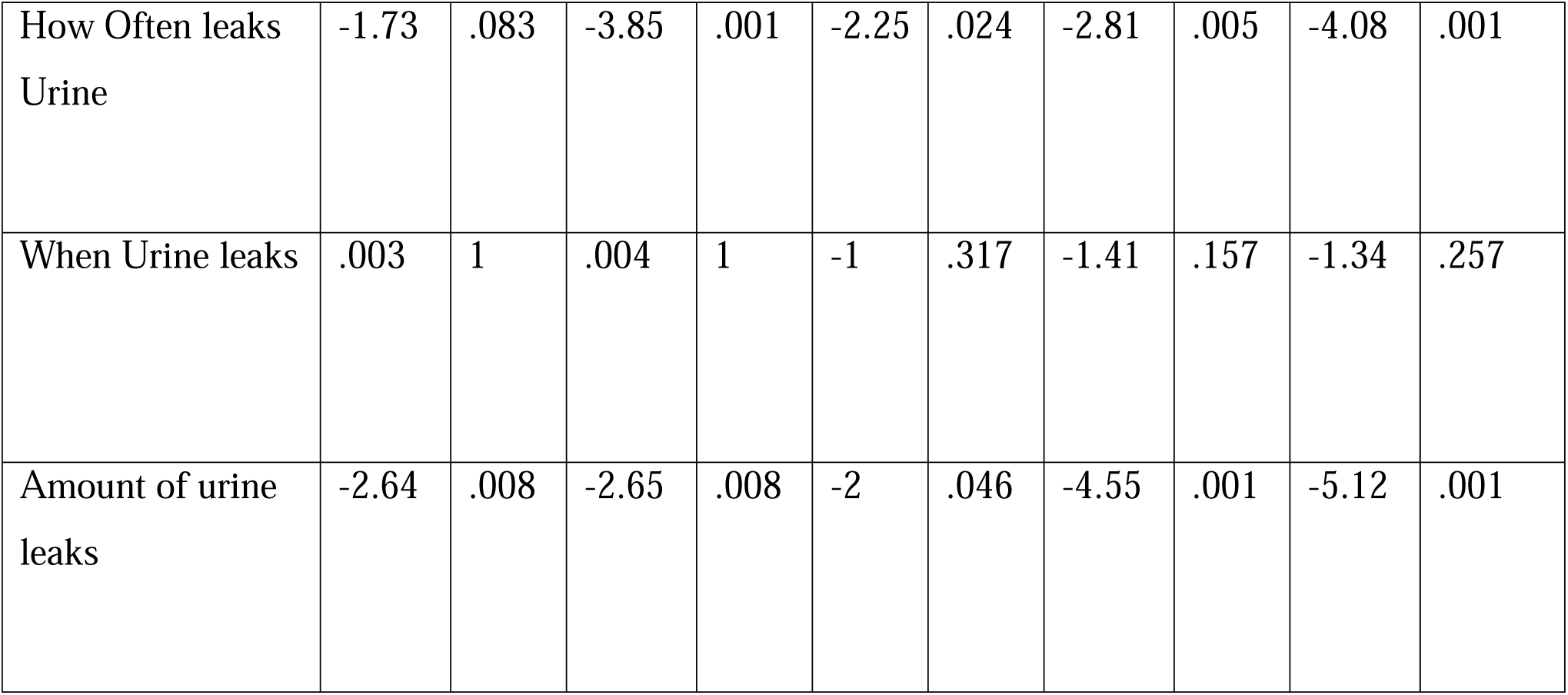
Improve of urine leakage (Week wise comparison)

**Table -3:** Improve of urine leakage
|  | Pretest to<br>W-1 |  | W-1 to W-2 |  | W-2 to W-3 |  | W-3 to W-4 |  | Pretest- W-4 |  |
| --- | --- | --- | --- | --- | --- | --- | --- | --- | --- | --- |
|  | Z | P | Z | P | Z | P | Z | P | Z | P |
| How Often leaks<br>Urine | -1.73 | .083 | -3.85 | .001 | -2.25 | .024 | -2.81 | .005 | -4.08 | .001 |
| When Urine leaks | .003 | 1 | .004 | 1 | -1 | .317 | -1.41 | .157 | -1.34 | .257 |
| Amount of urine<br>leaks | -2.64 | .008 | -2.65 | .008 | -2 | .046 | -4.55 | .001 | -5.12 | .001 |

## 4. Discussion

The aim of the study was to determine if maximum repetition of pelvic stabilization exercise impacts upon the management of stress urinary incontinence in female. The Specific objectives were to explore socio-demographics related to SUI and to observe the impact of maximum repetition of pelvic floor stabilization exercise upon pelvic floor strength & endurance, transverse abdominis strength and functional disability in stress urinary incontinence patients. Also, researchers observed the changes as per repeated measurement weekly in 4 weeks. Similar study by Bo et al. [17] had the aim of this article is to give an overview of the exercise science related to pelvic floor muscle (PFM) strength training, and to assess the effect of PFM exercises to treat stress urinary incontinence (SUI). Sixteen articles addressing the effect of PFM exercise alone on SUI were compiled by computerized search or found in other review articles. Kegel’s suggestion [18] was to perform 3–500 PFM contractions per day. However, suggestions for effective strength training from the exercise science are 8–12 contractions in three series 3–4 times a week for 15–20 weeks or more. Frequency of training varies between 10 repetitions every waking hour to half an hour 3 days a week. Holding periods vary between 2 and 3 s and 30–40 s. Exercise periods vary between 3 weeks and 6 months. Only a few research groups have used methods to measure PFM strength that were reproducible and valid. Statistically significant strength increase has been found after PFM exercise lasting from 3 to 6 months. Self-reported cure and success rates vary between 17% and 84%. Statistically significant improvement has been demonstrated on self-grading instruments, urethral closure pressure during cough, resting urethral pressure, functional urethral profile length, leakage episodes and pad tests with standardized bladder volume. The results of the long-term studies are promising. It is therefore concluded that PFM exercises are effective in treating SUI and to be more effective, PFM exercise has to be thoroughly taught and performed with weekly or monthly follow-up.

Another examination analyzed the viability of encouraging pelvic floor practices with utilization of bladder-sphincter biofeedback contrasted with preparing with verbal criticism dependent on vaginal palpation in 24 ladies with stress urinary incontinence. Verbal input preparing comprised of educating the patient to press the vaginal muscles around the inspector’s fingers and furnishing her with verbal execution criticism. Biofeedback patients got visual criticism of bladder pressure, stomach (rectal) weight, and outer butt-centric sphincter action. The biofeedback bunch improved the strength and specific control of pelvic floor muscles; the verbal input bunch didn’t. The two gatherings altogether decreased the recurrence of incontinence. The biofeedback bunch found the middle value of 75.9% decrease in incontinence, altogether more prominent than the 51.0% decrease appeared by the verbal criticism gathering. 12 out of 13 patients in the biofeedback treatment group improved by 60% or better out of the aim. Six patients in the verbal input bunch improved by 68% or better, and five were under 30% improved [19].

A Randomized Controlled Trial in ladies with stress urinary incontinence proposes that the Pelvic floor Muscle Training and Extracorporeal Magnetic Innervation are powerful in improving the pressure urinary incontinence and personal satisfaction in women. Another randomized control study recommended better results with a joined preparing of PFMT and Transversus Abdominis muscle than with PFMT alone in patients with stress urinary incontinence. Nonetheless, the preparation was more successful in the gathering of ladies who had lesser than three vaginal births [20].

An examination analyzing the preparation boundary for reinforcing the pelvic floor found the best convention to comprises of advanced palpation joined with biofeedback observing and vaginal cones, including multi week preparing boundaries, and ten reiterations for every arrangement in various positions [21].

## Limitation of the study

In spite of having the positive effect of maximum repetition of pelvic floor stabilization exercise in stress urinary incontinence, the study has some limitations. The study was a quasi-experimental study design with a single group and no control group. Thus, the study effect may not be suggested as an absolute effect. There were sources of bias, as there was no absolute blinding to the patients and therapists. The sample size was another limitation; a bigger sample size might have more absolute results.

## 5. Conclusion & Recommendations

Pelvic floor exercise with increasing repetition is an effective approach to improve stress urinary incontinence in females. The study found a significant impact on incontinence frequency, amount and associated quality of life for women with stress urinary incontinence after application pelvic floor exercise with maximum repetition. The improvement of Kegel exercise has been proven effective in the total duration time and separately in every week. The second and third week had more improvement in the incontinence questionnaire scale.

The study found the maximum 25 repetitions of exercise is effective, so further studies starting from 30 repetition and increasing duration up to 6 weeks are recommended. Hence, for future studies concentrating on the recommendations are encouraged: Randomized Control Study with parallel groups; female with stress, urge and mixed urinary incontinence; interventions with absolute treatment effect calculation.

## Data Availability

All data produced in the present study are available upon reasonable request to the authors

https://www.kaggle.com/kmamranhossain/urinary-incontinence

## Funding

No source of funding.

### Abbreviation

BHPl: Bangladesh Health Professions Institute
BMRC: Bangladesh Medical Research Council
CRP: Centre for the Rehabilitation of the Paralysed
ICIQ: International Classification of incontinence questionnaire
IRB: Institution Review Board
MMT: Manual Muscle Testing
SUI: Stress Urinary Incontinence
WHO: World Health Organization

